# Laboratory-Developed Test for SARS-CoV-2 Using Saliva Samples at the University of California, Riverside

**DOI:** 10.1101/2021.02.21.21251691

**Authors:** Rong Hai, Matthew Collin, Sophia Tsau, Daniel Raygoza, Juliet Morrison, Alexander J. Carrillo, Logan A. Collier, Walter Bayubay, Kenneth Han, Isgouhi Kaloshian, Katherine A. Borkovich

## Abstract

Here we describe a relatively quick, simple, economical and accurate laboratory developed test (LDT) for detection of SARS-CoV-2 in heated and diluted saliva samples without RNA extraction. Our protocol is a variation of the University of Illinois Urbana-Champaign SHIELD LDT. Differences include chilling of the samples during dilution and using a reduced volume for the qRT-PCR reactions. The level of detection for our LDT is 3125 copies/ml, which compares favorably with other saliva-based tests. Initial validation studies with a limited number of patient samples have demonstrated excellent agreement between results using our LDT and those obtained from external laboratories. The cost of consumables for our test is under $8 and a throughput of 1000 tests/day can be achieved with 3-4 personnel.

## INTRODUCTION

Several FDA-approved, commercially available diagnostic tests use reverse transcription combined with the polymerase chain reaction (RT-PCR). The University of California Riverside (UCR) Clinical Laboratory Improvement Amendments (CLIA) Extension Laboratory is using a modified version of the Laboratory Developed Test (LDT) established by the University of Illinois, Urbana-Champaign (UIUC) **(1)**. This test detects the presence of three viral genes: N-gene, S-gene and ORF1ab, with spiked MS2 phage as a sample control. The qRT-PCR is performed using the “TaqPath COVID-19 Combo Kit” marketed by Thermo Fisher Scientific (EUA200010/A001) **(2)** as described in the UIUC LDT. This decision was made in order to 1) maximize the biosafety practice; 2) simplify the experimental procedure, while retaining reasonable sensitivity, and 3) ensure a ready supply of test reagents, plasticware and other consumables.

Because we introduced several modifications to the UIUC protocol, including chilling samples during the dilution step and using a different volume for the qRT-PCR reactions, this modification is an LDT as described by CMS **(3)** and the FDA **(4)**. Thus, we are required to establish analytical and clinical validity of our adaptation of the test through experiments determining our limit of detection and through bridging assays comparing our results to those of other authorized testing sites.

### We consider our test to have the following advantages

1. We reduce cost and make the supply chain more sustainable by using a lower volume of Master Mix (5µl) for the qRT-PCR step.
2. Our test uses readily available consumables and is high throughput. We are utilizing 2-ml tubes for saliva collection that fit directly into our equipment. Multiple automated liquid handling robots are used for sample transfers.
3. The original saliva sample tube is not opened until after the heating step and inactivation of the virus, thus ensuring safety of the operators.
4. We chill samples after heating and during the dilution step, in order to better preserve the viral RNA.

## MATERIALS AND METHODS

### Establishment of a CLIA Extension Laboratory in the Multidisciplinary Research Building

We partnered with the Veitch Student Health Services High Complexity Laboratory at UCR to establish a CLIA Extension Laboratory at a non-contiguous location on campus, the Multidisciplinary Research Building (MRB). This was made possible by the FDA’s March 16^th^ Policy for Diagnostic Tests for Coronavirus Disease-2019 during the Public Health Emergency that streamlined approval or testing methods and work streams, and by Executive Order N-25-20 issued by California Governor Newsom that altered licensing requirements for laboratory personnel running diagnostic tests for SARS-CoV-2. We received approval of our application to establish a CLIA Extension Laboratory in MRB from the Centers for Medicare & Medicaid Services (CMS) and the California Department of Public Health (CDPH) on June 10, 2020. Our test has been validated in compliance with CMS CLIA regulations, but has not been submitted to the Food and Drug Administration (FDA) for Emergency Use Authorization.

Because we are not using the saliva samples for research, and are an extension of the clinical laboratory in Veitch Student Health Services, the UCR Institutional Biosafety Committee in the UCR Office of Research and Economic Development did not require a Biological Use Authorization or Institutional Review Board approval for research with human subjects. The Standard Operating Procedures (SOPs) for our testing protocol and our requirements for Personal Protective Equipment (PPE) were approved by the Veitch Student Health Services High Complexity Laboratory Director and the Assistant Biosafety Officer and High Containment Laboratory Director from UCR Environmental Health and Safety. All laboratory personnel were trained according to the approved SOPs and registered with CDPH. All personnel received HIPAA training. Using only barcoded samples with no protected health information on the tubes or on the laboratory computers ensured HIPAA compliance in the laboratory.

### Assay Development

As mentioned above, our assay is based on the University of Illinois Urbana-Champaign SHIELD LDT for detection of SARS-CoV-2 in saliva samples **(1)**.

#### Sample Collection

Each individual being sampled is provided with a fact sheet that describes the test being performed, possible results and the limitations of the test. They are also given a consent form to sign. They then generate a barcode label using a QR code on their phone and their ID. The individual is then given the barcode label, a 2-ml collection tube and a transfer pipet. The individual withdraws pooled saliva (not sputum) from inside the bottom cheek using the sterile transfer pipet. A volume of at least 0.5ml is transferred to the 2-ml barcoded tube.

The tube is securely capped and placed inside a secondary container. The transfer pipet containing excess saliva is discarded in a waste bag. Samples are then transported to the CLIA extension lab for testing.

#### Accessioning

Personnel #1, in full PPE, moves the container with the specimen tubes to the biosafety cabinet (BSC). Personnel #1 cleans the surface of each tube with an antiviral medical wipe and holds it so that Personnel #2 (in full PPE) can scan the barcode into the “Specimen Intake Log” file accessible on the computer. Samples are rejected if they have illegible or missing labels, significantly less volume than 0.5ml or large amounts of food particles. Personnel #2 notes the failed samples and Personnel #1 discards the failed samples in the waste container in the BSC. Cleaned tubes in 96-place racks are either stored in a secondary container at 4°C or transferred directly into the heating protocol. Providers are contacted to report failed specimens.

#### Heating and Dilution of Samples

Batches of up to 92 samples are removed by Personnel wearing full PPE from 4°C storage post-intake processing and transferred to a 95°C 96-place heating block in a dry bath in the BSC. Samples are heated for 30 minutes. Heating inactivates the virus and components of saliva that inhibit downstream PCR. Heated samples are removed from the heating block and cooled on ice or using chilled metal beads with places held by using 4 Opentrons OT-2 rack lids in a container. Samples are then transferred to 4 Opentrons OT-2 racks containing ice or chilled metal beads in the BSC. Caps are removed, making sure not to touch the rim of neighboring tubes, and discarded in the medical waste container in the BSC.

Sample racks are moved to the OT-2 in the BSC using a secondary container and placed into their positions. A 96-well plate with each well containing a 2x solution of TBE/Tween-20 is placed in a chilling rack in the robot. The Dilution/Aliquoting program is run, transferring 50µl of each heated and cooled saliva sample into a well containing 50µl of a 2x solution of TBE/Tween-20. The final concentration of the 1x solution with saliva is 89 mM Tris-Borate, 2 mM EDTA, pH 8.3 and 0.5% Tween-20 detergent. The robot pipets each solution up and down to mix the buffer into the saliva. The Dilution/Aliquoting program then pauses, and a plate with each well containing qRT-PCR master mix is placed in a chilling rack in the OT-2 (see below).

The 2-ml tubes containing the leftover heated saliva samples are removed from the OT-2 and placed in a 96-place covered rack and stored at 4°C until results are obtained. At that point, samples are discarded in a medical waste container.

#### qRT-PCR

Master Mix is prepared on ice or in a chilling rack using 0.5µl TaqPath RT-PCR COVID-19 Kit (Thermo Fisher Scientific, A47817) **(2)**, 2µl TaqPath 1-step (Thermo Fischer Scientific, A28523), 1.5 µl nuclease-free water and 1µl of MS2 phage for each sample. A multichannel pipet is used to dispense 5µl of master mix to each well of a PCR plate on ice or a chilling rack. Plates are used immediately. The plate is brought to the OT-2 liquid handler in the BSC that contains the heated and diluted saliva sample plate. The Dilution/Aliquoting protocol is then resumed, transferring 5µl of each diluted sample to a position in the 96-well PCR plate (plate is kept on ice or in a chilling rack). Negative and positive controls are then added to wells A1 and A12, respectively. PCR plates are sealed and transferred immediately to the qRT-PCR step. After completion of the Dilution/Aliquoting program, the 96-well plates containing the leftover diluted samples are sealed and placed at −80°C for long-term storage.

The 96-well RT-qPCR plate is then transferred to the Applied Biosystems Quant Studio 5 thermocycler. The RT-qPCR is run using the standard mode, consisting of a hold stage at 25°C for 2 min, 53°C for 10 min, and 95°C for 2 min, followed by 40 cycles of a PCR stage at 95°C for 3 sec then 60°C for 30 sec; with a 1.6°C/sec ramp up and ramp down rate.

The EDS output file produced during each qRT-PCR run is analyzed using the QuantStudio Design and Analysis software (version 1.2). The EDS file is then analyzed using the Applied Biosystems COVID-19 Interpretative Software and a CSV-formatted report generated. The software determines the results and validity of all tests, using predetermined cut-offs (see **Table 1**). The EDS file produced during the qRT-PCR run is then opened and the amplification plots for wells are compared to the CSV report. The amplification plots for the wells containing negative and positive controls and those producing invalid, inconclusive or positive results are visually checked for abnormalities. If an abnormality is observed, the result is discussed with the Laboratory Director to decide the next step. The CSV report file is then submitted to Student Health Services for reporting to patients and public health authorities.

**Table 1.** LDT Results Interpretation. Data from qRT-PCR were processed using QuantStudio Design and Analysis Software (version 1.2) followed by analysis using the Applied Biosystems COVID-19 Interpretative Software. Table is modified from Table 2 in the Thermo-Fisher TaqPath COVID-19 Combo Kit Instructions for Use **(2)**.

| ORF1ab | N gene | S gene | MS2 | Status | Result |
| --- | --- | --- | --- | --- | --- |
| NEG | NEG | NEG | NEG | INVALID | NA |
| NEG | NEG | NEG | POS | VALID | SARS-CoV-2 Not detected |
| Only one SARS-CoV-2 target = POS |  |  | POS or NEG | VALID | SARS-CoV-2 Inconclusive |
| Two or more SARS-CoV-2 targets = POS |  |  | POS or NEG | VALID | Positive SARS-CoV-2 |

## RESULTS/ASSAY VALIDATION

### Reduced Reaction Volumes

To minimize the overall cost of our assay, and to limit reagent use in the face of potential shortages, we reduced the quantity of reagents used for the RT-qPCR assay. After preliminary experiments showed that the test performed stably after such a reduction, we validated our workflow using 10µl total volume (5µl master mix and 5µl heated and diluted sample). We report the assay validation data conducted with the reduced volume protocol.

### LDT Validation

Validating an LDT requires measuring specific FDA/CDC-defined metrics and meeting or exceeding benchmarks. In the case of our SARS-CoV-2 LDT, these metrics were:

i. Measuring the assay limit of detection (LOD);
ii. Assessing clinical and analytical validity by running mock positive and negative samples at known concentrations,
iii. Obtaining saliva and nasal swab samples from SARS-CoV-2-positive and -negative individuals, with swabs submitted for independent analysis at local clinical diagnostic testing facilities and saliva analyzed using our LDT at UCR.

### Limit of Detection (LOD)

FDA policy recommends identifying the LOD of a molecular diagnostic by conducting a dilution series of SARS-CoV-2 RNA (or inactivated virus) in an artificial or real clinical matrix **(4)**. This guidance recommends that “laboratories test a dilution series of three replicates per concentration, and then confirm the final concentration with 20 replicates.” For the purposes of an EUA, the FDA defines the limit of detection as “the lowest concentration at which 19/20 replicates are positive.” In this section we describe the experimental data that establish the LOD of our LDT.

To experimentally determine our limit of detection, two different operators performed serial dilutions of SARS-CoV-2 positive control RNA with 5 replicates each, using 25 × 10^3^, 12.5 × 10^3^, 6250, 3125 and 1562.5 copies/ml **(Fig. 1)**. Each sample was spiked with bacteriophage MS2 as a control. At each dilution, we determined the Ct (cycle threshold) of three SARS-CoV-2 genes (N-gene, S-gene and ORF1ab) and the MS2 control. The Ct value is inversely proportional to the amount of starting target RNA, and represents the PCR cycle number at which the fluorescent signal of the reaction crosses a set threshold. This threshold was set manually based on background fluorescence of the reaction mixture.

**Figure 1.**
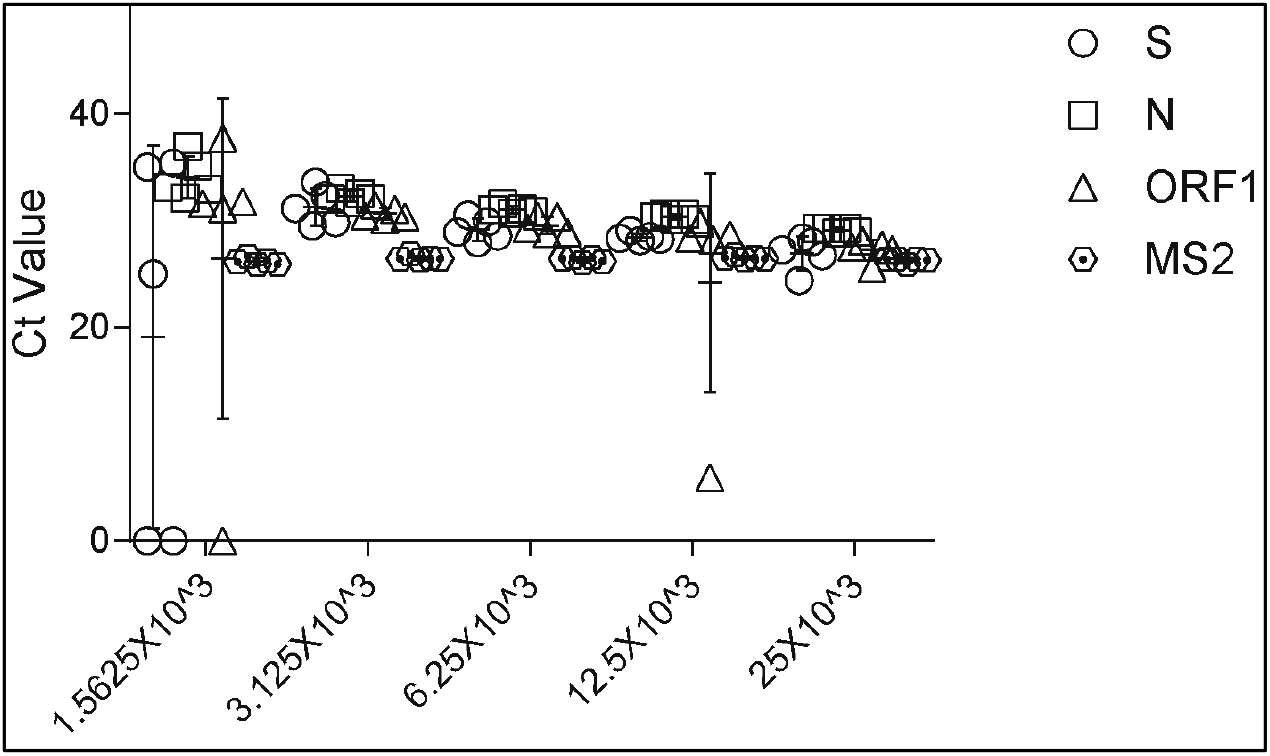
Limit of detection assay results. Positive control SARS-CoV-2 RNA from the Thermo-Fisher kit was diluted serially using saliva from negative donors that had been heated and diluted using TBE-Tween using our protocol. Samples from dilutions containing the indicated concentration of viral RNAs were combined with master mix and subjected to qRT-PCR according to the Assay Description. Ct values for the three viral genes (N, S and ORF1ab) and the MS2 bacteriophage control were plotted.

At our lowest concentration tested, 1562.5 copies/ml, we observed that the S gene was not amplified in two replicates and that ORF1ab was not amplified in one replicate. However, we detected amplification of all three SARS-CoV-2 genes at the four highest concentrations tested, 3125, 6250, 12.5 × 10^3^, and 25 × 10^3^ copies/ml. As expected, the MS2 control was amplified in all samples. **Based on these results, the limit of detection of our assay is 3125 viral genome copies per milliliter**.

### Reproducibility

We tested the reproducibility of the two lower concentrations used for the LOD study (1562.5 and 3125 genomic copies/ml) by performing the assay using 20 total technical replicates at each concentration **(Fig. 2)**. Samples were spiked with bacteriophage MS2 as a control.

**Figure 2.**
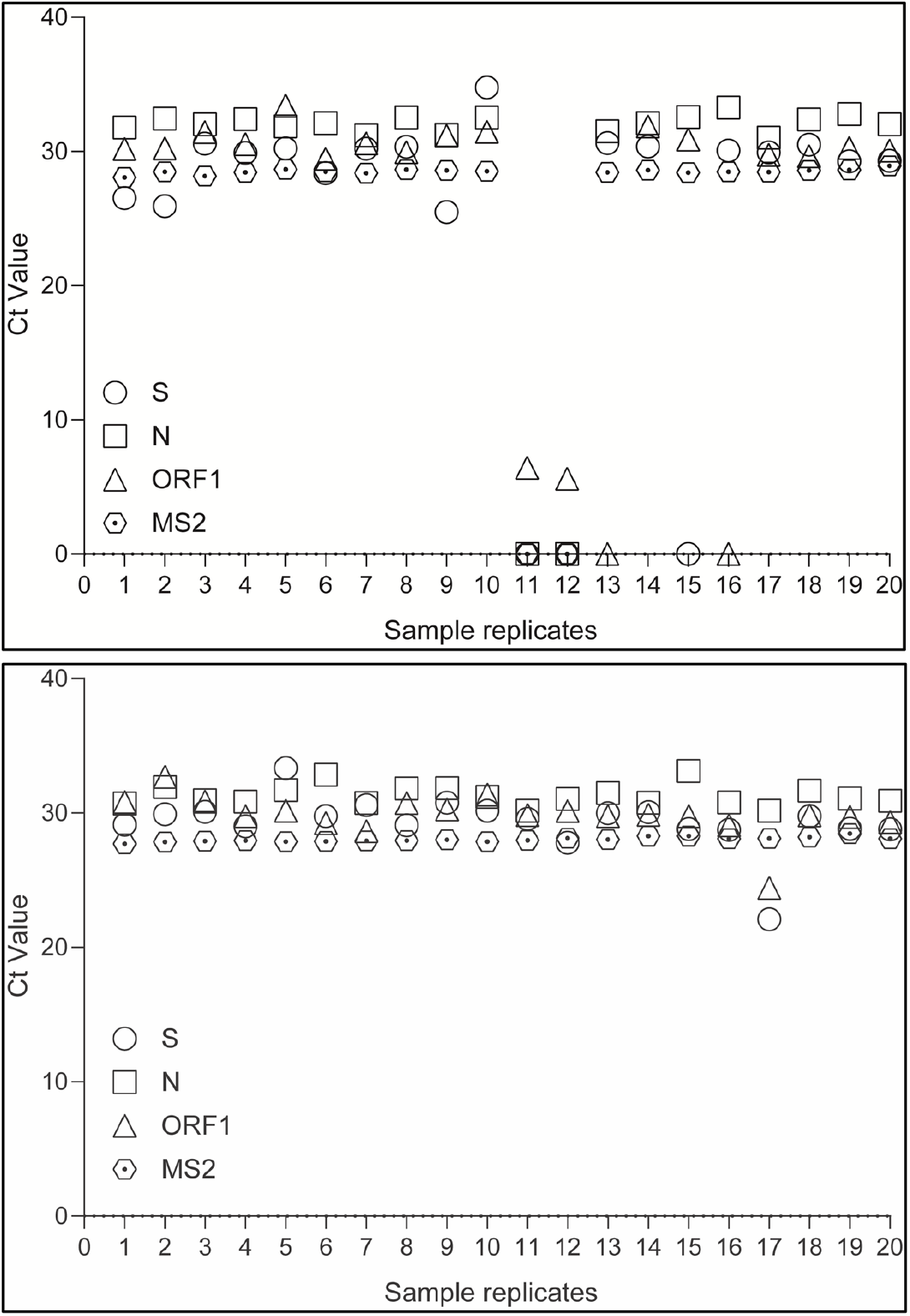
Reproducibility analysis results. Positive control SARS-CoV-2 RNA from the Thermo-Fisher kit was diluted to 1562.5 copies of each RNA/ml (**TOP**) or 3125 copies of each RNA/ml (**BOTTOM**) using saliva from negative donors that had been heated and diluted using our protocol. Samples were combined with master mix and subjected to qRT-PCR. Ct values for the three viral genes (N, S and ORF1ab) and the MS2 bacteriophage control were plotted.

Three of the 1562.5 genomic copies/ml replicates failed to amplify one of the three viral genes. In contrast, we detected all three SARS-CoV-2 genes, as demonstrated by consistent, positive Ct values, in the 3125 genomic copies/ml samples. **This is exceeding the required 19 out of 20 as defined by the FDA, and establishes 3125 viral genome copies per milliliter as our LOD**.

Our LOD of 3215 copies/ml compares favorably with other LDTs and EUAs for qRT-PCR detection of SARS-CoV-2 in saliva samples without RNA extraction. For example, the LOD for the UIUC SHIELD LDT is 1000 copies/ml **(1)**, while the LOD for the Yale SalivaDirect EUA is 6000-12,000 copies/ml **(5)**.

### Nonspecific Amplification

To meet the clinical validity criterion, 100% of all non-spiked saliva samples must be negative. To control for nonspecific amplification, we ran presumed negative saliva samples using our protocol (**Fig. 3**).

**Figure 3:**
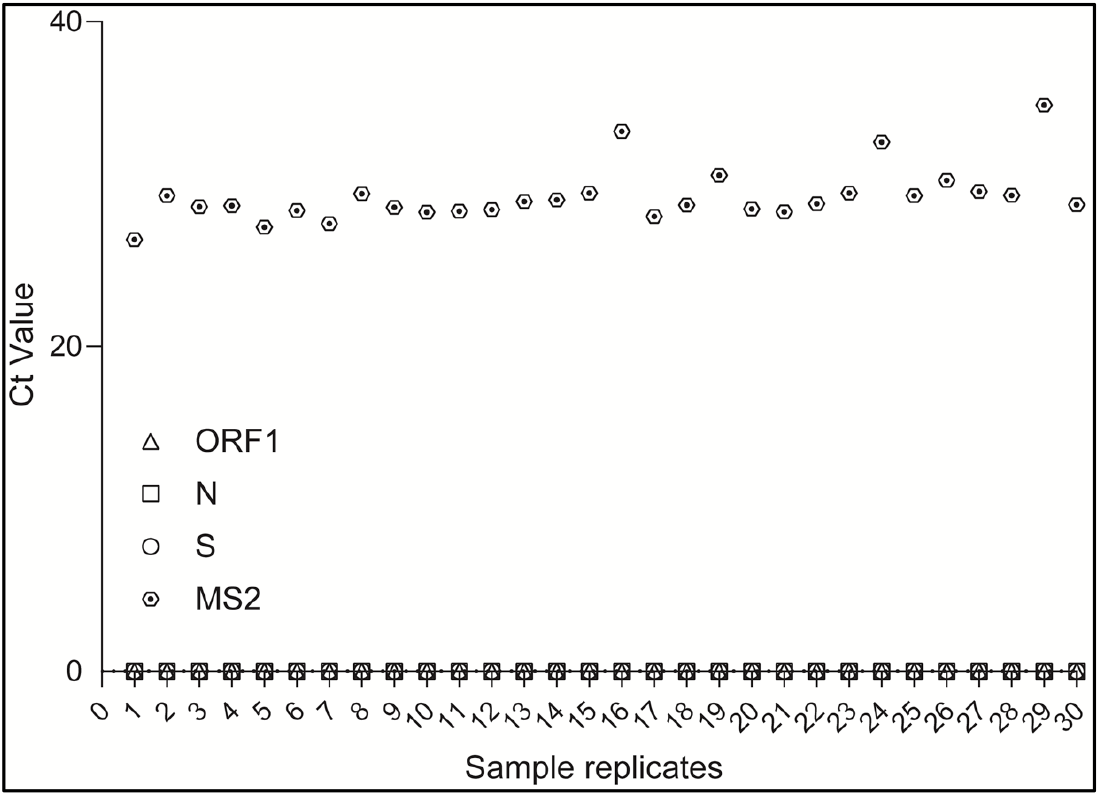
Negative Controls. Saliva samples from presumed negative donors were heated and diluted with TBE-Tween using our protocol. The samples were subjected to qRT-PCR according to the Assay Description, including spiked-in MS2 phage. Ct values for the three viral genes (N, S and ORF1ab) and the MS2 bacteriophage control were plotted.

MS2 yielded the expected Ct values for the 30 replicates, with no detectable amplification of any SARS-CoV-2 genes in any of the MS2-spiked saliva samples **(Fig. 3)**. The SARS-CoV-2 positive control from Thermo Fisher exhibited the expected amplification of all three SARS-CoV-2 genes, and the absence of MS2 phage (data not shown). This result demonstrates that non-specific amplification of SARS-CoV-2 genes was not observed in 30/30 samples, supporting a low false-positive rate for our LDT.

### Interpretation of Results

We are using the Thermo-Fisher TaqPath COVID-19 Combo Kit for qRT-PCR, as done previously for SHIELD at UIUC **(1)**. Therefore, we are using the parameters established for interpretation of results using nasal pharyngeal swabs/RNA extraction by Thermo-Fisher for their EUA **(Table 1) (2)**. Samples with no amplification of the three viral genes or MS2 are scored as invalid and a new sample obtained for testing. Samples with amplification of MS2 but not the three viral genes are scored as SARS-CoV-2 Not detected (Negative). Samples with amplification of only one viral gene with or without MS2 amplification are scored as SARS-CoV-2 Inconclusive and a new sample obtained for testing. Samples with two or more viral genes detected with or without MS2 amplification are scored as Positive SARS-CoV-2 (Positive).

### Clinical Validation Assay

We further validated our assay and workflow by submitting nasal swab or saliva samples from patients with positive and negative saliva test results for analysis at a different testing facility **(Table 2)**.

**Table 2.** Comparison of results from the UCR CLIA Extension Laboratory with those from external laboratories. Samples from 18 patients (ID# 1-18) were tested by the UCR CLIA Extension laboratory (Saliva/PCR) and an external testing laboratory (Nasal Swab/PCR or Saliva/Antigen). *The sample analyzed by the UCR laboratory was collected a week after the nasal swab sample was obtained for analysis by the outside laboratory.

| Sample ID# | UCR Saliva Test Result | External Lab |  | Pass/Fail |
| --- | --- | --- | --- | --- |
|  |  | Result | Test Type |  |
| 1 | POS | POS | Nasal Swab/PCR | Pass |
| 2 | POS | POS | Nasal Swab/PCR | Pass |
| 3 | POS | POS | Nasal Swab/PCR | Pass |
| 4 | POS | POS | Nasal Swab/PCR | Pass |
| 5 | POS | POS | Nasal Swab/PCR | Pass |
| 6 | POS | POS | Nasal Swab/PCR | Pass |
| 7 | Inconclusive/Retest* | POS | Nasal Swab/PCR | Inconclusive* |
| 8 | NEG | NEG | Nasal Swab/PCR | Pass |
| 9 | NEG | NEG | Nasal Swab/PCR | Pass |
| 10 | NEG | NEG | Nasal Swab/PCR | Pass |
| 11 | NEG | NEG | Saliva/Antigen | Pass |
| 12 | NEG | NEG | Nasal Swab/PCR | Pass |
| 13 | NEG | NEG | Nasal Swab/PCR | Pass |
| 14 | NEG | NEG | Nasal Swab/PCR | Pass |
| 15 | NEG | NEG | Nasal Swab/PCR | Pass |
| 16 | NEG | NEG | Nasal Swab/PCR | Pass |
| 17 | NEG | NEG | Nasal Swab/PCR | Pass |
| 18 | NEG | NEG | Nasal Swab/PCR | Pass |

The results for 17/18 clinical samples exactly matched those obtained for saliva samples using our LDT (94.4% agreement). Ct values for the known positive specimens spanned a large range (data not shown), supporting the dynamic range of our assay for clinical validation. Sample ID#7 was inconclusive according to results from the UCR testing laboratory and positive using a nasal swab sample at an external laboratory—it should be noted that this patient was recovering from COVID symptoms and a week had elapsed after the nasal swab sample was taken before the saliva sample was obtained and tested. Thus, it is possible that the viral load had decreased during the intervening week, leading to an inconclusive saliva test result from our LDT.

## Data Availability

All data referred to in the manuscript is available upon request.

## ACKNOWLEDGMENTS

This work was funded by the Office of Research and Economic Development at the University of California, Riverside. We thank members of the Innovative Genomics Institute SARS-CoV-2 Testing Consortium at UC Berkeley and Jeremy Sanford at UC Santa Cruz for advice early during development of our SARS-CoV-2 testing protocols. We thank numerous UCR faculty members who loaned equipment to the testing lab. We acknowledge Tran Phan from UCR Environmental Health and Safety and Jane Ngo-Trieu from UCR Student Health Services for support and safety training. We thank Sabrina Schuster, Sarah Acrey, Joann Braga, Jeanette Westbrook and Linda Roney for assistance with procurement of equipment and supplies. We acknowledge Jing Li, Josh Hoerger, Bart Kats and Paul Hackman for assistance with Information Technology and regulatory issues. We are indebted to Vice Chancellor of Research and Economic Development Rodolfo Torres, School of Medicine Dean Deborah Deas, College of Agricultural and Natural Sciences Dean Kathryn Uhrich, faculty in the Department of Microbiology and Plant Pathology, and many other faculty and staff on the UCR campus for providing advice, space and financial support during development of the CLIA Extension Laboratory and SARS-CoV-2 testing protocol.

## REFERENCES

1. Diana Rose E. Ranoa, Robin L. Holland, Fadi G. Alnaji, Kelsie J. Green, Leyi Wang, Christopher B. Brooke, Martin D. Burke, Timothy M. Fan, Paul J. Hergenrother. Saliva-Based Molecular Testing for SARS-CoV-2 that Bypasses RNA Extraction. bioRxiv 2020.06.18.159434; doi: https://doi.org/10.1101/2020.06.18.159434.

2. Thermo Fisher Scientific TaqPath COVID-19 Combo Kit. Instructions for Use. Publication Number MAN0019181, Revision D.0., May 6, 2020.

3. https://www.cms.gov/Regulations-and-Guidance/Legislation/CLIA/Downloads/Research-Testing-and-CLIA.pdf

4. Policy for Coronavirus Disease-2019 Tests During the Public Health Emergency (Revised). May 11, 2020. https://www.fda.gov/media/135659/download

5. Accelerated Emergency Use Authorization (EUA) Summary SARS-CoV-2 RT-PCR Assay. Yale School of Public Health, Department of Epidemiology of Microbial Diseases. https://www.fda.gov/media/141192/download

